# High Food Insecurity in Latinx Families and Associated COVID-19 Infection in the Greater Bay Area, California

**DOI:** 10.1101/2020.10.11.20210906

**Authors:** Milagro Escobar, Andrea DeCastro Mendez, Maria Romero Encinas, Janet M Wojcicki

**Affiliations:** Division of Pediatric Gastroenterology, Hepatology and Nutrition, University of California, San Francisco

## Abstract

**Background:** Food insecurity impacts nearly one-in-four Latinx households in the United States and has been exacerbated by the novel coronavirus or COVID-19 pandemic.

**Methods:** We examined the impact of COVID-19 on household and child food security in three preexisting, longitudinal, Latinx urban cohorts in the San Francisco Bay Area (N=375 households, 1,875 individuals). Households were initially recruited during pregnancy and postpartum at Zuckerberg San Francisco General Hospital (ZSFG) and UCSF Benioff prior to the COVID-19 pandemic. For this COVID sub-study, participants responded to a 15-minute telephonic interview. Participants answered 18 questions from the US Food Security Food Module (US HFSSM), described food consumption, housing and employment status, and history of COVID-19 infection as per community or hospital-based testing. Food security and insecurity levels were compared with prior year metrics.

**Results:** We found low levels of household food security in Latinx families (by cohort: 29.2%; 34.2%; 60.0%) and child food security (56.9%; 54.1%; 78.0%) with differences between cohorts explained by self-reported levels of education and employment status. Food security levels were much lower than those reported previously in two cohorts where data had been recorded from prior years. Reported history of COVID-19 infection in households was 4.8% (95% Confidence Interval (CI); 1.5-14.3%); 7.2% (95%CI; 3.6-13.9%) and 3.5% (95%CI; 1.7-7.2%) by cohort and was associated with food insecurity in the two larger cohorts (p=0.03; p=0.01 respectively).

**Conclusions:** Latinx families in the Bay Area with children are experiencing a sharp rise in food insecurity levels during the COVID-19 epidemic. Food insecurity, similar to other indices of poverty, is associated with increased risk for COVID-19 infection. Comprehensive interventions are needed to address food insecurity in Latinx populations and further studies are needed to better assess independent associations between household food insecurity, poor nutritional health and risk of COVID-19 infection.

## Background

Food insecurity is common among Latinx families,, the largest ethnic minority group in the United States and has been further exacerbated by the novel coronavirus (COVID-19) (Lauren et al., 2020). Prior to COVID-19, 1 in 9 households faced food insecurity with a higher prevalence in Latinx families (Coleman-Jensen et al., 2013; Coleman-Jensen et al., 2015). Latinx individuals comprise over 60 million individuals in the United States (Noe-Bustamante, 2010) and are twice as likely as White Americans to face poverty including aspects of food insecurity (Maani and Galea, 2020). Preliminary reports since the beginning of the COVID-19 shelter-in-place lockdowns in the United States (March 2020) suggest that Latinx families have had higher increases in food insecurity compared to Whites (32% versus 18%) with even higher rates in families with children (Schanzenbach and Pitts, 2020).

Higher rates of food insecurity in Latinx communities can be explained by sociodemographic and economic factors specific to these communities. Latinx individuals are more likely to be low-income, recent immigrants, have young children, live in larger households, and reside in urban areas that have been historically prone to food insecurities (Hernandez et al., 2013; Rabbitt et al., 2016; Potochnick and Arteaga, 2018; Van Hook and Balistreri, 2006). Furthermore, Latinx adults may have been most impacted by loss of employment due to sectors that were particularly impacted by COVID-19 shutdowns including the hotel industry, food services, healthcare, and manufacturing (Clark et al., 2020). Similar to other high-risk groups, Latinx families have relied on government food services since the start of the pandemic (Bermudez, 2020). However, the number of Latinx families served is likely underestimated as many Latinx immigrants who contribute to the US economy are not able or resistant to receiving economic relief in context of the COVID-19 pandemic due to lack of legal status. Additional disparities in Latinx communities include crowded living conditions, housing insecurity, education gaps, few family resources, all which create higher risk for COVID-19 infection (Bixler et al, 2020).

We have previously assessed the prevalence of food insecurity in Latinx low-income Latinx families residing in six counties of the San Francisco Bay Area finding high rates prior to COVID-19 (Nagata et al., 2018). Slightly more than half of the households previously surveyed had high food security (58.3%) whereas the rest experienced marginal food security (8.6%), low food security (28.2%), and very low food security (4.9%). Food insecurity in children has been linked to lower academic performance, anxiety, obesity and diabetes. Adults with food insecurity can have lower scores on mental and physical health tasks, higher risk of depression and anxiety, chronic disease, and poorer overall health status. Previous studies have also suggested that food insecurity may increase risk for infectious disease such as HIV when exposed due to poor immunological health (Rollins, 2007)

To our understanding, no study has investigated the impact of COVID-19 on food security in a longitudinal, Latinx sample of participants residing in an urban U.S. content. We examined food security during COVID-19 using three prospective longitudinal studies of Latinx households in the 6 counties of the San Francisco Bay Area. We evaluated levels of food security and insecurity during COVID-19 compared with previous levels (prior to COVID-19) in these populations as well as the association between COVID-19 infection and food insecurity.

## Methods

### Recruitment and Consent

Mothers were recruited from three previous longitudinal birth cohort studies: the Hispanic Eating and Nutrition Study (HEN) (n=201) (Kjaer et al., 2018; Wojcicki et al., 2011), Latinx, Eating, and Diabetes Study (LEAD) (n=97) (Ville et al., 2017; Wojcicki et al., 2018) and the Telomeres At Birth Study (TAB) (n=424). Participants for both HEN and LEAD were recruited during pregnancy as previously described and TAB participants were recruited post-partum before being discharged from the hospital. All participants from HEN, LEAD and TAB have been followed up over the course of approximately 13, 8, and 1-2 years respectively. These cohorts were initially recruited to evaluate the relationship between prenatal and early postnatal exposures and development of childhood obesity. Inclusion criteria for HEN and LEAD included self-identifying as Latinx and expecting a healthy newborn infant as previously described and participants were primarily recruited from Zuckerberg San Francisco General Hospital (ZSFG) with a smaller percentage from UCSF Benioff for HEN (Wojcicki et al., 2011; Wojcicki et al., 2017). We initially recruited all race/ethnic backgrounds for TAB but restricted entry criteria to Latinx as a part of a second phase of our study. Seventy-point two percent of participants from TAB were recruited at UCSF Benioff and the remained at ZSFG.

To assess participation in the COVID-19 food insecurity sub-study, participants were contacted telephonically during the months of May to September 2020 using previous contact information. Participants were asked for their language preference (English or Spanish) and were provided information about the COVID-19 sub-study by bilingual research coordinators and the study principal investigator. For those participants who were interested, we obtained verbal consent and participants took part in a 15-minute interview. Food insecurity was assessed using the US Household Food Security Scale Module (US HFSSM) that we have previously used annually with HEN and LEAD studies to assess food insecurity (USDA, 2020). We additionally asked questions about COVID-19 symptoms and testing, housing and employment status and food consumption. Participants were compensated for their time for participation with a $15 gift card to Target or Amazon based on participant preference. The principal investigator of the study (JW) and four research coordinators including (ME, ADC and MR) conducted all interviews. The Committee on Human Research (CHR), the UCSF Institutional Review Board approved all aspects of the study (IRB numbers 11-06334; 16-06163; 16-18535).

### Procedures

Food insecurity was measured using the 18-item US Household Food Security Scale Module. The timeframe of the questions was modified to collect data from when COVID-19 lockdowns began (March 2020) to present time of interview. The Food Insecurity Module including questions regarding adult and children’s eating habits (USDA, 2020). Scoring was done automatically by REDCap, with higher scores indicative of higher level of food insecurity. Following previously established food insecurity categories the following categories were determined based on number of affirmative responses: high food security or food secure (0 affirmative responses), marginal food security (1–2 affirmative responses), low food security (3– 7 affirmative responses), and very low food security (8–18 affirmative responses). Child food security was defined as high food security or food secure (0 affirmative responses), marginal food security (1 affirmative response), low food security (2-4 affirmative response) and very low food security (5-8 affirmative responses). We compared mean ± SD food security scores and categorical markers of food security/insecurity with prior year assessments of food security/insecurity in two out of three cohorts (HEN and LEAD). We had no prior assessment of food security/insecurity in TAB.

Food assistance information was collected prior to the COVID-19 pandemic as part of our previous longitudinal data collection (self-reported participation in the Special Supplemental Program for Women, Infants and Children’s program (WIC) or participation in the Supplemental Nutrition Assistance Program (SNAP; previously known as food stamps)). Similarly, maternal education level (primary or less than a high school diploma, high school diploma, some college or more) and primary language use (Spanish, English or bilingual) was also collected at study entry for all participants. As part of the COVID-19 sub-study we asked about adult household employment status, number of total household members as well as crowding indices including size of house (number of bathrooms and bedrooms).

Participants were asked about COVID-19 symptoms, laboratory-based testing as well as duration of symptoms for all household members since middle of March 2020 until time of interview.

### Statistical Analysis

Household and child food security levels and corresponding insecurity were calculated including mean score ± standard deviation (SD) as well as percentages by category. Data was stratified by cohort using Stata 15.0. Levels of household and child food security were assessed for normality using graphical assessments as well as statistical tests to assess normality including Shapiro-Wilk. As there were departures from normality, non-parametric statistical tests were used to assess associations including Wilcoxon rank sum, Kruskal-Wallis and Spearman rank order correlation coefficient for continuous predictors and chi-squared tests for categorical ones. Associations between household and child food insecurity levels (continuous measures and categorical variables) and education level (high school over lower versus more than high school education), language use (Spanish, English or bilingual), COVID-19 testing positivity, and employment status (unemployed or adult household employment) were assessed using non-parametric methods. Multivariable logistic models were used to evaluate independent predictors of household COVID infection in relation to food insecurity in TAB and HEN cohorts adjusting for potential confounders such as unemployment and crowding that were significant at p<0.05 in bivariate analysis.

## Results

We recruited 200 families from TAB, 111 from HEN and 64 from LEAD for a total participation of 375 families. Mean household size was 4.8±1.7 for TAB, 5.3±2.0 for LEAD, and 4.9±1.6 for HEN (Table 1). Total sample size for all three longitudinal studies combined was 1,875. All studies reported a comparable number of children in households with a mean of 2.4±1.1 for TAB, 2.7±1.1 for LEAD and 2.9±1.2 for HEN (Table 1).

**Table 1:** Demographics and Household Specifics by Bay Area Cohort.

| <u>Cohort Name</u> | **Telomeres at Birth (TAB)**<br>**% (N)**<br>**Mean ±SD**<br>**N=200** | **Latinx, Eating and Diabetes (LEAD)**<br>**% (N)**<br>**Mean ±SD**<br>**N=65** | **Hispanic, Eating and Nutrition (HEN)**<br>**% (N)**<br>**Mean ±SD**<br>**N=111** |
| --- | --- | --- | --- |
| <u>Household size</u> |  |  |  |
| No. of people | 4.8±1.7 | 5.3±2.0 | 4.9±1.6 |
| No. of children | 2.4±1.1 | 2.7±1.1 | 2.9±1.2 |
| Number share Bedroom | 2.5±1.0 | 2.8±1.3 | 2.3±1.1 |
| Number share bathroom | 3.3±1.6 | 4.5±1.6 | 3.8±1.6 |
| <u>Food Assistance</u> |  |  |  |
| SNAP/Food Stamps | Not Collected | 47.1% | 43.4% |
| WIC | 41.4% | 15.7% | 72.9% |
| <u>Language Use</u> |  |  |  |
| Spanish | 38.7% | 90.5% | 91.1% |
| English | 56.8% | 4.2% | 8.9% |
| Bilingual (English/Spanish) | 4.5% | 4.8% | --- |
| <u>Maternal Education</u> |  |  |  |
| Highest Education Level |  |  |  |
| High School Or less | 34.3% | 79.4% | 75.2% |
| <u>Employment</u> |  |  |  |
| All household adults unemployed | 24.1% | 35.2% | 38.8% |

A relatively high percentage of HEN and LEAD participants reported receiving SNAP or food stamps prior to the onset of the COVID-19 pandemic (43.4% and 47.1% respectively). We did not have this information on TAB participants. All of the HEN and LEAD participants self-identified as Latinx with 80.6% of the TAB participants identifying as Latinx, 13.8% non-Hispanic Caucasian and 5.3% as Asian including Pacific Islander. We only included those TAB participants in this COVID-19 sub-study who identified as Latinx (n=200).

A high percentage of HEN and LEAD participants reported using Spanish as a primary language (91.1% and 90.5%) with a much lower percentage using Spanish (38.7%) for TAB participants. Other sociodemographic differences between HEN, LEAD and TAB included higher maternal education levels for TAB. In TAB a low 34.3% had a high school education or less (65.7% with university or postgraduate qualifications) compared with 79.4% of LEAD with high school as the highest qualification (20.6% with university/postgraduate) and 75.2% for HEN (24.8% with university/postgraduate). In the COVID-19 era, a relatively high percentage of households had no adults who were currently employed, 24.1% for TAB, 35.2% of LEAD and 38.8% of HEN.

### Food Security

A low percentage of two out of three cohorts report household food security with 29.2% for LEAD and 34.2% for HEN being food secure. A much higher (60.0%) report household food security for TAB participants (Table 2). Similarly, a high percentage of participants in HEN and LEAD report low and very low food security (46.2% for LEAD and 49.5% for HEN) with a much lower 26.5% for TAB. Additionally, slightly more than half of participants from HEN and LEAD report child food security. LEAD and HEN reported low levels of child food security (56.9% and 54.1% respectively) with a much higher 78.0% for TAB (Table 2).

**Table 2:** Levels of Household, Child Food Security and COVD-19 Infection by Bay Area Cohort.

|  | **Telomeres at Birth (TAB)**<br>**% (N)**<br>**Mean ±SD**<br>**N=200** | **Latinx, Eating and Diabetes (LEAD)**<br>**% (N)**<br>**Mean ±SD**<br>**N=65** | **Hispanic, Eating and Nutrition (HEN)**<br>**% (N)**<br>**Mean ±SD**<br>**N=111** |
| --- | --- | --- | --- |
| <u>Levels of Household</u> |  |  |  |
| <u>Food Security</u> |  |  |  |
| Food Security Score, Mean ±SD | 1.6±2.7 | 3.3±3.7 | 3.5±3.6 |
| High Food Security (Food Secure) | 60.0 (120) | 29.2 (19) | 34.2 (38) |
| Marginal Food Security | 13.5 (27) | 24.6 (16) | 16.2 (18) |
| Low Food Security | 20.5 (41) | 33.9 (22) | 31.5 (35) |
| Very Low Food Security | 6.0 (12) | 12.3 (8) | 18.0 (20) |
| <u>Levels of Child</u> |  |  |  |
| <u>Food Security</u> |  |  |  |
| High Food Security (Food Secure) | 78.0 (156) | 56.9 (37) | 54.1 (60) |
| Marginal Food Security | 8.0 (16) | 10.8 (7) | 10.8 (12) |
| Low Food Security | 12.0 (24) | 26.2 (17) | 32.4 (36) |
| Very Low Food Security | 4 (2.0) | 6.2 (4) | 2.7 (3) |
| <u>COVID Infection History</u> |  |  |  |
| (positive test March-Sept 2020) | 7/200 (3.5%)<br>(95%CI; 1.7-7.2%) | 3/62 (4.8%)<br>(95%CI; 1.5-14.3%) | 8/111(7.2%)<br>(95%CI; 3.6-13.9%) |

### Associations between Food Insecurity and Socio-demographics

There were no associations between socio-demographics including maternal education level and employment status with household or child food security in HEN or LEAD cohorts. Higher household crowding factors as indicated by more people sharing a bedroom or bathroom were associated with having less food security in HEN but not LEAD cohorts (Rho=0.29, p<0.01 for number sharing a bedroom for total food security score and Rho=0.28, p<0.01 for number sharing a bath; Rho=0.33, p<0.01 for number sharing a bedroom for child food security score and Rho=0.27, p<0.01 for number sharing a bathroom).

However, sociodemographic variables associated with poverty including not being employed and having a lower education level (high school versus any university or postgraduate training) were associated with increased levels of household and child food insecurity in TAB (p<0.01). Furthermore, crowding metrics (persons/bedroom and persons/bathroom) were also significantly associated with household and child food insecurity in TAB (p<0.01). Additionally, being a Spanish language speaker was also highly correlated with having less household and child food security in TAB (z=-6.8 p<0.01 household security and z=-4.2, p<0.01 for child security) but not in HEN or LEAD.

### Change from Food Insecurity in Prior Years

Prior data from these same cohorts in the last year and prior years suggest that there is much more severe food insecurity with fewer families now being food secure with COVID-19. In LEAD interviews from one year ago, 76.9% of families were household food secure and 87.9% had child food security. In the 6 months prior to COVID, we assessed at sub-sample of HEN families (n=38) and 79.0% had household security and 89.2% had child food security. A larger study of HEN participants assessed greater than 5 years ago (n=161) found that many of these families had struggled with food security in prior years with 56.6% with household food security and 69.3% with child food security.

### Association with COVID

There were 8 households surveyed in HEN that had COVID-19 infections as indicated by self-report positive reverse transcription polymerase chain reaction (RT-PCR) tests out of the 111 total households surveyed that also had food security reported (7.2%, 95%CI; 3.6-13.9%) (Table 2). Of these, only 12.5% had household food security compared with 34.7% of those households without individuals with COVID-19 positive members (p=0.03). Adjusting for crowding factors including persons per bedroom and bathroom, food insecurity category was not associated with COVID-19 positivity (p=0.60). The percentage of positive households was lower among LEAD (3 out of 62 households having a COVID-19 infection or 4.8% (95%CI; 1.5-14.3%) (Table 2). There was no association between COVID-19 infection and household food insecurity in LEAD. Similarly, the infection rate in TAB household was lower than HEN (7 out of 200 or 3.5% (95%CI; 1.7-7.2%)) (Table 2). There was also a trend towards reduced food security among TAB households that were infected (42.9% food secure versus 60.6% of those uninfected and 28.6% had the highest level of food insecurity compared with 5.2% of those uninfected, p=0.01). In a model adjusting for employment status, crowding as indicated by household members sharing a bathroom and Spanish language use, increased household food insecurity score trended towards a positive association with household COVID infection (OR 1.25, 95%CI 0.97-1.62; p=0.088).

## Discussion

Racial and ethnic minorities historically have worse outcomes during and after disasters as they already face a range of economic, political, and social barriers (Hutchins et al., 2009; Méndez et al., 2020). These include but are not limited to racial discrimination, exploitation, limited English-speaking proficiency, and lack of legal status (Méndez et al., 2020).

The overall prevalence of food insecurity in California between 2017-2019 was 9.9% having low food insecurity and 3.6% with very low food security (USDA, 2019). Overall national levels of food insecurity were higher during this time period for Hispanic household (15.6%).

Food insecurity has increased for American children and adults since the start of COVID-19 pandemic with elevations in every state (Schanzenbach and Pitts, 2020). Data from the Census Pulse Household Survey suggest that overall food insecurity tripled from 9.4% to 29.5% for households with children during COVID-19. Similarly, we report dramatic increases in food insecurity in our low income Latinx cohorts in the San Francisco. These population groups already had low rates of food security. Prior to COVID-19, LEAD families reported having 76.9% food security but post-COVID-19 only 38.1% report food security. Similarly, HEN food security declined from 79.6% of adults having food security to 34.2%. Decreases in child food security were also significant with 87.9% secure prior to COVID declining 56.9% in LEAD and 89.2% to 54.1% in HEN. Our results in HEN and LEAD correspond with the high food insecurity rates (only 36% with food security) reported by US adults living <250% below the federal poverty line during the COVID-19 pandemic by Wolfson and Leung (2020).

Our findings of high food insecurity among three Latinx cohorts the San Francisco Bay Area suggest rapid growth of food insecurity in urban areas impacted by COVID-19 shutdowns. Latinx individuals are among the most likely to be pushed into poverty as a result of pandemic-related unemployment (Schanzenbach and Pitts, 2020; Feeding America, 2020). A significant percentage of our participants did not have any adult household members with employment at the time of the interview (more than one third of HEN and LEAD households and a quarter of TAB), and unemployment is strongly associated with food insecurity (Abrams et al., 2019; Schaznenbach and Pitts, 2020). Additionally, Latinx individuals, such as the individuals in cohorts, who faced food insecurity prior to COVID-19 likely have more severe food insecurity post-pandemic and are particularly at risk for adverse outcomes (Wolfson and Leung, 2020).

Common coping strategies for food insecurity prior to COVID-19 include use of food pantries, borrowing money, going to homes of friends or family for meals, or sending children to relatives (Gundersen and Ziliak, 2015). Some, if not all of these tactics to buffer families against food insecurity, are more limited given the recommended physical distancing measures being taken to prevent COVID-19 transmission (Wasserman et al, 2020). Even when food pantry resources are available, Latinx compared with other groups may face language barriers or concerns about legal status which limit use (Wright et al., 2020). Shame can also result in people hiding hunger rather than going to food pantries and further increasing food vulnerability (Zepeda, 2018).

### Differences Among Latinx Cohorts

There were large differences between our Latinx cohorts in levels of food insecurity. Part of this can be explained by socio-demographic differences by cohorts. HEN and LEAD were recruited primarily at Zuckerberg San Francisco General Hospital, the local safety net county hospital with TAB recruited primarily at UCSF Benioff, a tertiary care academic hospital. The women recruited for HEN and LEAD cohorts reported a low percentage with any education beyond high school at recruitment (20.6% for LEAD and 26.1% for HEN) versus a much higher 65.7% for TAB. Low education status is the greatest predictor of unemployment (van Zon et al., 2017) and since the emergence of COVID-19, employment losses are associated with low education levels (Kochhar, 2020). Higher rates of household and child food insecurity in HEN and LEAD versus a much lower level in TAB (60% with high food security) parallel the lower education levels and potential earning power in these cohorts (Abrams et al., 2020; Schanzenbach & Pitts, 2020).

Additionally, over 90% of mothers from the HEN and LEAD were Spanish-speakers while 65% of the TAB mothers were primary English speaking. Being fluent in English leads to greater job opportunities (Park, 1999) also potentially contributing to food insecurity (Abrams et al., 2020; Schanzenbach and Pitts, 2020). Our previous studies with HEN and LEAD have also found that the majority of the adults in these households are recent immigrants and foreign born (Wojcicki et al., 2011; Ville et al., 2017; Wojcicki et al., 2018) potentially places these families at a greater disadvantage in terms of accessing food and other government resources (Page et al., 2020).

There was little difference in reported crowding indices (the number sharing bedrooms and bathrooms) between the cohorts and although there were only associations between these metrics and food insecurity in TAB and HEN possibly based on differences in sample size. We did not see any differences in food insecurity and education level or employment status for LEAD and HEN possibly because there was little variation in education level with the majority not having any advanced education beyond high school. We did not assess for underemployment common among Latinx adults but HEN and LEAD may have higher levels of underemployment (Golden and Kim, 2020). Meanwhile for TAB, the association between education level and employment status and food insecurity was highly significant. Those with less education have greater food insecurity, possibly due to the heterogeneity in education levels for this population.

### Necessary Interventions for Latinx Families

Our data suggest the urgent need for interventions for Latinx families with children, particularly those who are Spanish-speaking and with low education and employment levels in the context of the COVID-19 crisis. Data from our TAB cohort points to the heterogeneity among Latinx families in the Bay Area and the potential need for targeted interventions in specific communities. Due to urgent need, some safety net clinics in California have had creative responses in terms of bringing food to families. There needs to be more funding and more development of these efforts (Ugwu-Oju, 2020; Center for Care Innovations, 2020). Furthermore, individuals from underrepresented groups tend to not have enough accessible information in their preferred language and mode of communication (DeBruin et al., 2012). For instance, 29% of Latinx in the United States have no English fluency (US Department of Health and Human Services, Office of Minority Health, 2020). There can also be distrust of the government and health agencies due to poor history and absence cultural competency (DeBruin et al., 2012; Page et al., 2020). The best implementations for COVID-19 relief will be made through the inclusivity of cultural richness, such as having information in multiple languages, modes of communication, and a diverse workforce to implement these interventions and work with diverse communities.

### Risk for COVID-19 Infection with Household Food Insecurity

Previous researchers have noted an increased risk for COVID-19 morbidity and mortality due to low quality nutrition, obesity and associated metabolic disease (Belanger et al., 2020). While crowding factors and poverty are associated with risk for COVID-19 infection and food insecurity, food insecurity may be independently associated with increased risk for COVID-19 infection through poorer immune health and increased susceptibility to disease. Previous studies, particularly related to risk for HIV infection in the context of HIV exposure suggest that food insecurity may be an independent risk factor for infectious disease (Rollins, 2007). Other studies suggest that household food insecurity puts children in a more susceptible position for tuberculosis infection (Faust et al., 2018). In multivariable analyses adjusting for crowding, unemployment and Spanish language use in the TAB cohort, food insecurity score trended towards association with household COVID infection (OR 1.25, 95%CI 0.97-1.61). Further studies are needed with larger cohorts to assess the role between food insecurity, poor nutritional status and risk for infectious disease.

## Conclusions and Limitations

Further population-based studies are needed with larger cohorts and heterogeneous cohorts to assess the impacts of COVID-19 in other high-risk population groups, in addition to Latinx. Rural communities may face different food insecurity stressors and as such our findings may not be relevant to rural areas. Government and non-governmental agencies need to prioritize poverty-alleviating measures including mitigating food insecurity as a means to address the public health demands of COVID-19 infection.

## Data Availability

The data is available based on requests to the principal investigator.

## Acknowledgements

This project was supported by the generosity of the Eric and Wendy Schmidt by recommendation of the Schmidt Futures program, through Covid Catalyst at the Center of Emerging and Neglected Diseases. Additional funding came from NIH NIDDK 080825 097458 as well as the Hellman Family Foundation and Allen Foundation and Marc and Lynn Benioff.

## References

Abrams SA, Avalos A, Gray M, Hawthorne KM. High Level of food insecurity among families with Children seeking routine care at federally qualified health centers during the Coronavirus disease 2019 pandemic. J Pediatr X. 2020;4:100044.

Bauer L. About 14 million children in the US are not getting enough to eat. Brookings. https://www.brookings.edu/blog/up-front/2020/07/09/about-14-million-children-in-the-us-are-not-getting-enough-to-eat/. Published 2020. Accessed September 28, 2020.

Belanger M, Hill M, Angelidi A, Dalamaga M, Sowers J, Mantzoros C. Covid-19 and Disparities in Nutrition and Obesity. New England Journal of Medicine. 2020;383(11):e69. doi:10.1056/nejmp2021264

Bermudez E. As demand for food skyrockets due to coronavirus, food banks play catch-up. LA Times. May 9 2020.

Bixler D, Miller AD, Mattison CP, et al SARS-CoV-2-associated deaths among persons aged <21 years - United States, February 12-July 31, 2020. MMWR Morb Mortal Wkly Rep. 2020;69(37):1324–1329

Center for Care Innovations. LA Clinics Respond to Food Insecurity During COVID-19. https://www.careinnovations.org/resources/la-clinics-respond-to-food-insecurity-during-covid-19/, accessed October 6, 2020.

Clark E, Fredricks K, Woc-Colburn L, Bottazzi ME, Weatherhead J. Disproportionate impact of the COVID-19 pandemic on immigrant communities in the United States. PLoS Negl Trop Dis. 2020;14(7):e0008484.

Coleman-Jensen A, Gregory C, Singh A. Household food security in the United States in 2013. SSRN Electron J. 2014. doi:10.2139/ssrn.2504067

Coleman-Jensen A, Rabbitt MP, Gregory CA, Singh A. Household food security in the United States in 2015. Usda.gov. https://www.ers.usda.gov/webdocs/publications/79761/err215_summary.pdf?v=3774.6. Accessed September 23, 2020.

Curran MA, Minoff E. Supporting children and families through the pandemic, and after: The case for a US child allowance. Social Sciences & Humanities Open. 2020;2(1):100040.

DeBruin D, Liaschenko J, Marshall MF. Social justice in pandemic preparedness. Am J Public Health. 2012;102(4):586–591.

Duncan B, Hotz V, Trejo S. Hispanics in the U.S. Labor Market. In National Research Council (US) Panel on Hispanics in the United States; Tienda M, Mitchell F, editors. Hispanics and the Future of America. Washington (DC): National Academic Press. Ncbi.nlm.nih.gov. https://www.ncbi.nlm.nih.gov/books/NBK19908/. Published 2006. Accessed September 28, 2020.

Faust L, McCarthy A and Y Schreiber. Recommendations for the Screening of Latent Tuberculosis Infection in Indigenous Communities: A systematic Review of Screening Strategies Among High-Risk Groups in Low-Incidence Countries. BMC Public Health 2018; 198; 979.

Feeding America. Feedingamerica.org. The impact of the Coronavirus on food insecurity. https://www.feedingamerica.org/sites/default/files/2020-04/Brief_Impact%20of%20Covid%20on%20Food%20Insecurity%204.22%20%28002%29.pdf. Published 2020. Accessed September 27, 2020.

Golden L and J Kim. Underemployment Just Isn’t Working for US Part-time Workers. The Center for Law and Social Policy. https://www.clasp.org/publications/report/brief/underemployment-just-isnt-working-us-part-time-workers, accessed October 4, 2020.

Gundersen C, Ziliak JP. Food insecurity and health outcomes. Health Aff (Millwood). 2015;34(11):1830–1839.

Hernandez DC, Reesor L, Murillo R. Gender disparities in the food insecurity-overweight and food insecurity-obesity paradox among low-income older adults. J Acad Nutr Diet. 2017;117(7):1087–1096.

Hutchins SS, Fiscella K, Levine RS, Ompad DC, McDonald M. Protection of racial/ethnic minority populations during an influenza pandemic. Am J Public Health. 2009;99 Suppl 2(S2):S261–70.

Kjaer TW, Faurholt-Jepsen D, Medrano R, et al Higher birthweight and maternal pre-pregnancy BMI persist with obesity association at age 9 in high risk Latino children. J Immigr Minor Health. 2019;21(1):89–97.

Kochhar R. Hispanic women, immigrants, young adults, those with less education hit hardest by COVID-19 job losses. Pew Research Center. https://www.pewresearch.org/fact-tank/2020/06/09/hispanic-women-immigrants-young-adults-those-with-less-education-hit-hardest-by-covid-19-job-losses/. Published 2020. Accessed September 28, 2020.

Lauren BN, Silver ER, Faye AS, Woo Baidal JA, Ozanne EM, Hur C. Predictors of household food insecurity in the United States during the COVID-19 pandemic. bioRxiv. 2020:2020.06.10.20122275. doi:10.1101/2020.06.10.20122275

Maani N, Galea S. COVID-19 and underinvestment in the health of the US population. Milbank Q. 2020;98(2):239–249.

Méndez M, Flores-Haro G, Zucker L. The (in)visible victims of disaster: Understanding the vulnerability of undocumented Latino/a and indigenous immigrants. Geoforum. 2020;116:50–62.

Nagata JM, Gomberg S, Hagan MJ, Heyman MB, Wojcicki JM. Food insecurity is associated with maternal depression and child pervasive developmental symptoms in low-income Latino households. J Hunger Environ Nutr. 2019;14(4):526–539.

Noe-Bustamante L, Hugo-Lopez M and J Manuel Krogstad. US Hispanic Population Surpassed 60 Mission in 2019, but growth has slowed. Pew Research. 2109 https://www.pewresearch.org/fact-tank/2020/07/07/u-s-hispanic-population-surpassed-60-million-in-2019-but-growth-has-slowed/

Page KR, Venkataramani M, Beyrer C, Polk S. Undocumented US. immigrants and covid-19. N Engl J Med. 2020;382(21):e62

Park J. The Earnings of Immigrants in the United States: The Effect of English-Speaking Ability. The American Journal of Economics and Sociology. 1999;58(1):43–56. https://www-jstor-org.ucsf.idm.oclc.org/stable/3487874?seq=1#metadata_info_tab_contents. Accessed September 28, 2020.

Potochnick S, Arteaga I. A decade of analysis: Household food insecurity among low-income immigrant children. J Fam Issues. 2018;39(2):527–551.

Rabbitt M, Smith MD, Coleman-Jensen A. Food Security among Hispanic Adults in the United States, 2011-2014. United States Department of Agriculture, Economic Research Service; 2016.

Rabbit MP, Smith MD and Coleman-Jensn A. Food insecurity and Hispanic diversity. Usda.gov. https://www.ers.usda.gov/amber-waves/2016/july/food-insecurity-and-hispanic-diversity. Accessed September 18, 2020

Rollins N. Food Insecurity-A Risk Factor for HIV Infection. PLOS Medicine October 23, 2007. https://doi.org/10.1371/journal.pmed.0040301.

Schanzenbach D, Pitts A. How much has food insecurity risen? Evidence from the Census Household Pulse Survey [Institute for Policy Research (IPR) Rapid Research Report]. Northwestern Institute for Policy Research. June 10, 2020. Accessed September 26, 2020. https://www.ipr.northwestern.edu/documents/reports/ipr-rapid-research-reports-pulse-hh-data-10-june-2020.pdf

Schhneider B, Martinez S, Ownes A. Barriers to Educational Opportunities for Hispanics in the United States. Ncbi.nlm.nih.gov. https://www.ncbi.nlm.nih.gov/books/NBK19909/. Published 2006. Accessed September 28, 2020.

Schwarzenberg SJ, Georgieff MK, Committee on Nutrition. Advocacy for improving nutrition in the first 1000 days to support childhood development and adult health. Pediatrics. 2018;141(2). doi:10.1542/peds.2017-3716

Seligman HK, Laraia BA, Kushel MB. Food insecurity is associated with chronic disease among low-income NHANES participants. J Nutr. 2010;140(2):304–310.

Stuff JE, Casey PH, Szeto KL, et al Household food insecurity is associated with adult health status. J Nutr. 2004;134(9):2330–2335.

Ugwa-Oju D. How a pandemic exposed racial inequality, food insecurity in California’s Central Valley. Fresno Bee. https://www.fresnobee.com/fresnoland/article243240666.html, accessed October 6, 2020.

United Nations Food and Agriculture Organization. Trade Reforms and Food Security Rome, Italy: Food and Agriculture Organization;2003.

USDA Key statistics & graphics, Food Security Status of US Households in 2019. Usda.gov. https://www.ers.usda.gov/topics/food-nutrition-assistance/food-security-in-the-us/key-statistics-graphics.aspx. Accessed September 27, 2020.

USDA Economic Research Service. Survey Tools. 2020 Usda.gov. https://www.ers.usda.gov/topics/food-nutrition-assistance/food-security-in-the-us/survey-tools/. Accessed September 28, 2020.

United States Department of Health and Human Services. Office of Minority Health. Profile: Hispanic/Latino Americans. https://minorityhealth.hhs.gov/omh/browse.aspx?lvl=3&lvlid=64, Accessed October 11, 2020.

van Hook J, Balistreri KS. Ineligible parents, eligible children: Food Stamps receipt, allotments, and food insecurity among children of immigrants. Soc Sci Res. 2006;35(1):228–251.

van Zon S, Reijneveld S, Mendes de Leon C, Bültmann U. The impact of low education and poor health on unemployment varies by work life stage. Int J Public Health. 2017;62(9):997–1006. doi:10.1007/s00038-017-0972-7.

Ville AP, Heyman MB, Medrano R and JM Wojcicki. Early Antibiotic Exposure and Risk of Childhood Obesity in Latinos. Childhood Obesity 2017; 13(2): https://doi.org/10.1089/chi.2016.0235.

Wasserman D, van der Gaag R, Wise J. The term “physical distancing” is recommended rather than “social distancing” during the COVID-19 pandemic for reducing feelings of rejection among people with mental health problems. Eur Psychiatry. 2020;63(1):e52.

Wojcicki JM, Holbrook K, Lustig RH, Epel E, Caughey AB, Munoz RF, Shiboski SC and MB Heyman. Chronic Maternal Depression is Associated with Reduced Wieght Gain in Latino Infants from Birth to 2 Years. PLOS One 2011; 6(2): e16737.

Wojcicki JM, Medrano R, Lin J, Epel E. Increased Cellular Aging by 3 Years of Age in Latino, Preschool Children Who Consume More Sugar-Sweetened Beverages. Child Obes 2018 Apr 1:14(3): 149–157.

Wolfson JA, Leung CW. Food insecurity and COVID-19: Disparities in early effects for US adults. Nutrients. 2020;12(6):1648

Wright KE, Lucero J, Crosbie E. “It’s nice to have a little bit of home, even if it’s just on your plate” – perceived barriers for Latinos accessing food pantries. J Hunger Environ Nutr. 2020;15(4):496–513.

Zepeda L. Hiding hunger: food insecurity in middle America. Agric Human Values. 2018;35(1):243–254.

